# Performance of Adenosine Deaminase assay in diagnosis of pulmonary & extrapulmonary tuberculosis

**DOI:** 10.1101/2021.04.30.21256025

**Authors:** Anuja Kawle, Arti Mishra, Vinita Hutke, Seema Shekhawat, Amit Nayak, Rima Biswas, Nitin Chandak, Vijay Agrawal, Hatim Daginawala, Lokendra Singh, Rajpal S. Kashyap

**Author notes:** Corresponding author: Dr. Rajpal S. Kashyap.

## Abstract

There has been a lot of debate about the use of the Adenosine Deaminase (ADA) for tuberculosis (TB) diagnosis. In the present study, to overcome this debate, we have planned a large scale study in all forms of TB to access the performance of ADA for diagnosis of TB. we have also studied the performance of the ADA in the Tb prognosis. ADA activity was evaluated using Guisti and Galanti method. The sensitivity of the ADA test is impressive in all forms of TB clinical samples analysed for the study (PTB [82%], TBM [85%] TBAR [85%]), and (ATB) [84%]. However, the specificity was variable in pulmonary TB, but satisfactory in extrapulmonary TB (EPTB) cases (i.e TBM [89%], (ATB) [88%], TBAR [88%]). In the follow-up samples, ADA value declined drastically after the anti TB medication (ATT). Our study, which consists of a large number of samples, suggests that the ADA has very limited value in the diagnosis of PTB, and hence it is not recommended for PTB diagnosis. We recommend, ADA test is quite useful for EPTB diagnosis along with other clinical correlations and in absence of other advanced diagnostic tests. However, on the other ADA can be used as a prognostic test for all forms of TB.

## 1. Introduction

Tuberculosis (TB) is a serious public health threat. Rapid and accurate diagnosis is the key to control TB, especially in high TB burden countries. The only available gold standard test for TB is the sputum smear microscopy and microbiological culturing, which after a certain limit is found to be low in sensitivity and time-consuming [1-3]. The low specificity of chest X-rays used for the diagnosis of smear-negative TB is another major concern for misdiagnosis [4]. There is no second doubt that with the increased incidence of pulmonary TB (PTB), the incidence of extra-pulmonary TB (EPTB) has also increased. According to a survey, it was found that the diagnosis of EPTB has always been a difficult task & requires further research [5]. During the past decade, various Molecular and Immunological methods have been developed and widely used for the diagnosis of PTB and EPTB. Although these methods are quite beneficial to some extent for diagnosis of all forms of TB, they suffer from low efficacies or unbelievably expensive laboratory diagnoses [6]. A number of studies have already been reported in the usefulness of ADA for diagnosis of PTB and EPTB and can be used as a supplementary or supporting test in suspected cases of TB where sophisticated laboratory facilities are not available [7,8]. The previous studies on ADA were reported in a small number of samples, hence it was difficult to conclude the advantages and limitations of the ADA test [9]. The major concern with small sample size studies is the interpretation of results where a chance of false-positive results or overestimation of the magnitude of study reliability of the ADA test is high. In the present study, we evaluated the performance of the ADA test in a large set of clinical samples of all forms of TB cases. In addition, to know whether the ADA test can be used as a prognostic marker or not, we have done three months follow-up study in TB patients who were on anti-TB treatment (ATT), and ADA activity in these clinical samples along with other parameters were evaluated.

## 2. Material and methods

### 2.1 Ethics Statement

The present observational study (prospective and retrospective) study was approved by the Institutional Ethics Committee, of Central India Institute of Medical Sciences (CIIMS), Nagpur. Written consents were taken from each participant after a detailed oral explanation about the study.

### 2.2 Study Design

The present study was conducted in a Tertiary care hospital from January 1, 2010, to December 31, 2018. A total of 7498 participants were enrolled in the study. The study participant was divided into two major groups, namely PTB (n= 4123) and EPTB (n=1512) based on their clinical symptoms. EPTB group was further divided into three groups these include Tuberculous meningitis (TBM), TB arthritis (TBAR) & Abdominal TB (ATB). Those participants who refuge to participate in later stages, who were less than <18 years of age, infection other than TB, patients without proper history, Insufficient sample for analysis, and those who expired were excluded (n=1863) from the study as shown in fig 1. A detailed history of all participants was taken. Serum, CSF, Synovial Fluid samples, and abdominal fluid samples were collected from the respective TB group for the required investigations in aseptic conditions. Samples were collected before the initiation of ATT and were stored at −20°C until they were used for experimental analysis.

**Figure 1:**
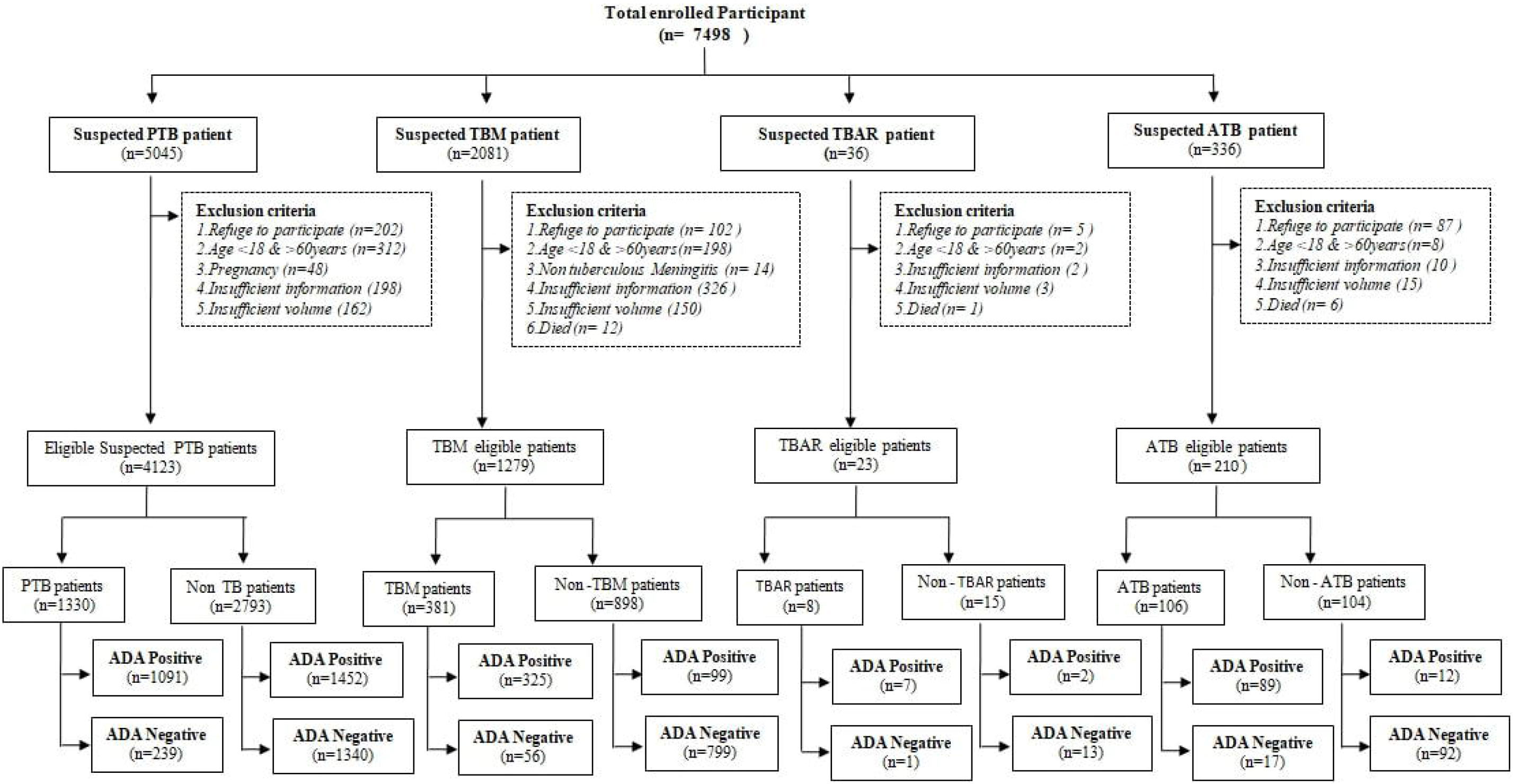
Flow chart showing a large scale study for evaluation of the performance of ADA assay for diagnosis of PTB, TBM, TBAR & ATB.

### 2.3 Patient and samples

#### 2.3.1 PTB group

Patients in the PTB group cases were classified into two different groups viz. PTB group and non-PTB group are based on their following clinical profile.

PTB patients (n=1330): Patients who were sputum culture positive were grouped in the PTB group. sputum microscopy was done on two serial sputum samples by staining with Ziehl Neelson Stain as per the guidelines of India’s Revised National Tuberculosis Control Program (RNTCP). Culture isolates of M.tb on Lowenstein-Jensen medium, obtained after 6 weeks were considered positive. In patients whose acid-fast bacilli (AFB) staining and culture test were negative, the diagnosis of PTB was done on the following clinical observations: 1) Low-grade fever for 2-3 weeks, 2) loss of appetite, 3) abnormal chest X-ray, 4) weight loss, 5) night sweats, 6) chronic cough with or without expectoration/hemoptysis; 7) chronic chest pain for 2-3 weeks, and 8) history of TB.

Non-PTB patients (n=2793): This included patients admitted to the hospital with non-tuberculous pulmonary infections such as renal & hepatic disorders, cancer bilateral bronchitis, bacterial infection, viral infections, non-specific fever, urinary tract infection, anemia, pneumonia, neurological disorders, gastrointestinal infections, malaria, PID, skin disease, COPD, and PUO. Healthy volunteers were also included in the study showing no apparent clinical impairment with normal chest radiographs.

Blood samples: 2ml venous blood was collected from all the patients of the PTB group. Blood was allowed to clot, and after centrifugation (1000× g, 10 min), serum was separated and stored at -20_ο_ C until use.

#### 2.3.2 EPTB group

TBM group (n=1279): It is classified into two groups viz. TBM patients (n=381) and non-TBM patients (n=898). The diagnostic observations for the above classification were based on the criteria as reported by Kashyap et.al [10].

CSF samples: CSF was collected by lumbar puncture of all the TBM group patient patients. Besides the routine microscopic and biochemical analysis, staining was done in all cases with gram stain, acid-fast stain, and India ink. Samples obtained from all the patients were stored at -20 °C until further use.

#### 2.3.3 TB arthritis group (TBAR)

TBAR patient ((n=8): Patient in this group was confirmed by the presence of M.tb in the synovial fluid by staining and/or culture. In conditions where the test showed negative results, the patients were then diagnosed by clinical symptoms such as Persistent knee ache, b. swelling in the knee, c. radiographs along with common TB symptoms such as fever & weight loss.

Non-TBAR patient (n=15): This included patients who were admitted to the hospital for acute or chronic defined, non-TB joint diseases including rheumatoid arthritis and gout. Synovial Fluid samples: For the collection of synovial fluid, the patient’s joint area was cleaned with iodine or similar solution. The needle was carefully inserted through the skin into the joint space. The fluid was then drawn through the needle into a sterile syringe. Samples obtained from all the patients were stored at -20 °C until further use.

#### 2.3.4 Abdominal tuberculosis (ATB) group

ATB patient (n=106): Diagnosis of ATB was made based on a positive culture of M.tb from the abdominal fluid (Ascitic fluid, Pleural Fluid, Peritoneal fluid). In absence of culture result the patients were diagnosed by clinical symptoms viz. a. Abdominal pain, b. Abdominal mass, c. Abdominal distension, d. Diarrhea, e. Histological findings along with common TB symptoms such as fever, loss of weight. In addition, biochemical findings were also studied, which showed elevated protein levels with decreasing glucose values and/or pleocytosis with lymphocytic predominance and ultrasound of the chest suggestive of ATB. Along with the radiographic features, the supporting clinical diagnosis was lung parenchymal infiltration mainly involving apical and/or mid zone, miliary shadows, and pleural effusion. Along with the above mentioned clinical features, any one of the radiological features was considered sufficient as supportive evidence.

Non-ATB group (n=104): This included patients admitted to the hospital for liver cirrhosis, chronic peritonitis, and acute colitis. Along with this, control group cases were also included with patients showing comorbidities such as pneumonia, lung cancer, bronchitis, respiratory sinus arrhythmia, and asthma.

ATB samples: For the collection of ascitic fluid, the patient was allowed to lie on his/her back with a head at 45°C–90°C elevation. The area where the needle was to be inserted was cleaned with iodine or a similar solution. The anesthetic was administered to numb the area. The paracentesis needle was carefully inserted into the abdomen. About 1000 to 1500 ml of fluid was removed. For diagnosis, 50 ml of the fluid was stored at -20°C until further use.

#### 2.3.5 Follow up samples

Patients were followed at regular intervals for three months. All patients were confirmed to have taken ATT. Normal health regained by all the patients was judged based upon clinical criteria, including improvements in cough, fever, appetite, weight gain as well as radiographic evidence.

## 3. ADA assay

ADA activity in all the collected samples was determined by the method of Guisti and Galanti^10^ based on the Berthelot’s reaction, which is the formation of the colored indophenol complex from ammonia liberated during deamination of adenosine. The development of color was quantified by spectrophotometer (Systronic, India). One unit of ADA is defined as the amount of enzyme required to release 1mMol of ammonia/min from adenosine at standard assay conditions. Results were expressed as units per liter per minute (U/L/min). A sample with a cut-off value of ≥15 U/L/min was considered as a positive test result.

## 4. Statistical analysis

All the statistical analysis was performed in Medcalc statistical software version 10. The receiver operating curve (ROC) was used to calculate the cut off values, sensitivity specificity ADA assay for diagnosis of PTB and EPTB. A comparison of the followup parameters and ADA level was done by t-test. A P value <0.05 was considered significant.

## 5. Results

A total of 7498 patients, which include 5045 patients of PTB, 2081patient of TBM, 36 patients of TBAR & 336 Patients of ATB, were enrolled in the study between January 1, 2010, to December 31, 2018. Out of the enrolled suspected patient for PTB, TBM, TBAR & ATB, only 5635 were found to be eligible for further study based on the predefined inclusion-exclusion criteria. These include 4123 were PTB, 1279 were TBM, 210 ATB patient & 23 TBAR patients. Out of the eligible suspected patients, 1330 (32 %) were diagnosed with active pulmonary TB according to WHO Guidelines [11, 12] 381 (29%) were diagnosed with TBM based on the criteria as reported by Kashyap et.al [10, 13] Similarly 8 (34 %) & 106 (50 %) patients were diagnosed with TBAR & ATB in respective suspected patient samples (Fig 1). The result of the ADA assay in the eligible suspected PTB, TBM, TBAR & ATB patients show the positivity of 82% (1091/1330), 85 % (325/381) 87.5% (7/8), & 84% (89/106) among the confirmed patient in the respective category. Similarly positivity of 52% (1452/2793), 11% (99/898), 13% (02/15) & 12% (12/104) was observed in Non-PTB, Non TBM, non TBAR, & non ATB patient respectively (Fig 1 and Table 1). Fig 2 (a-d) shows the box plots presentation of ADA activity in PTB, TBM, TBAR & ATB patients & respective non TB groups. The mean activity of ADA was significantly higher (p<0.05) in PTB (25.83 ± 4.90) TBM (5.72 ± 2.43), TBAR (18.17 ± 5.52), & ATB ((19.66 ±4.02) patients as compared to their respective non-PTB patients. Fig 3 represents the bar graph of the follow-up total leukocyte count (TLC), neutrophil count, Lymphocyte count, Protein, Sugar, CSF: blood glucose ratio, and ADA activity (U/dL) in the CSF samples of TBM patients (n=381) before & after ATT for three months. There is a significant (P<0.05) decrease in TLC (32.12 ± 11.93) neutrophil count (7.4 ± 3.6), Lymphocytes count (15.33 ± 7.14), Protein (43.41 ± 14.62), and ADA activity (6.11 ± 2.26) in the follow-up CSF samples of TBM patients (Fig 3 a-d & g), while increase (P < 0.05), in the CSF Sugar (52 ± 7.38), & CSF blood glucose ratio (0.47 ± 0.07), was observed after ATT for three months (Fig 3 e & f). Similarly, there is a decline (P < 0.05)) in the Total WBC count (6020± 1523), ESR (24.60 ± 8.66), and ADA activity (9.80 ± 2.52), in the serum samples of PTB patients (n=1330) after taking ATT for three months as shown in Fig 4 (a-c).

**Table 1:** Sensitivity (%) and Specificity (%) of ADA assay for the diagnosis of PTB, TBM, TBAR & ATB cases

| **TB Group** | **Sensitivity (%)** | **Specificity (%)** |
| --- | --- | --- |
| **PTB group (n=4123)** |  |  |
| Pulmonary TB (n= 1330) | 1091 (82%) | 239 (18%) |
| Non PTB (n=2793) | 1452 (52%) | 1340 (48%) |
| **EPTB group (n= 1512)** |  |  |
| TBM group (n= 1279) |  |  |
| TBM (n= 381) | 325 (85%) | 56 (15%) |
| Non TBM (n=898) | 99 (11%) | 799 (89%) |
| TBAR Group |  |  |
| TBAR (n=8) | 7 (85%) | 01 (15%) |
| Non TBAR (n=15) | 02 (12%) | 13 (88%) |
| ATB Group |  |  |
| ATB (n=106) | 89 (84%) | 17 (16%) |
| Non ATB (n=104) | 12 (12%) | 92 (88%) |

**Figure 2:**
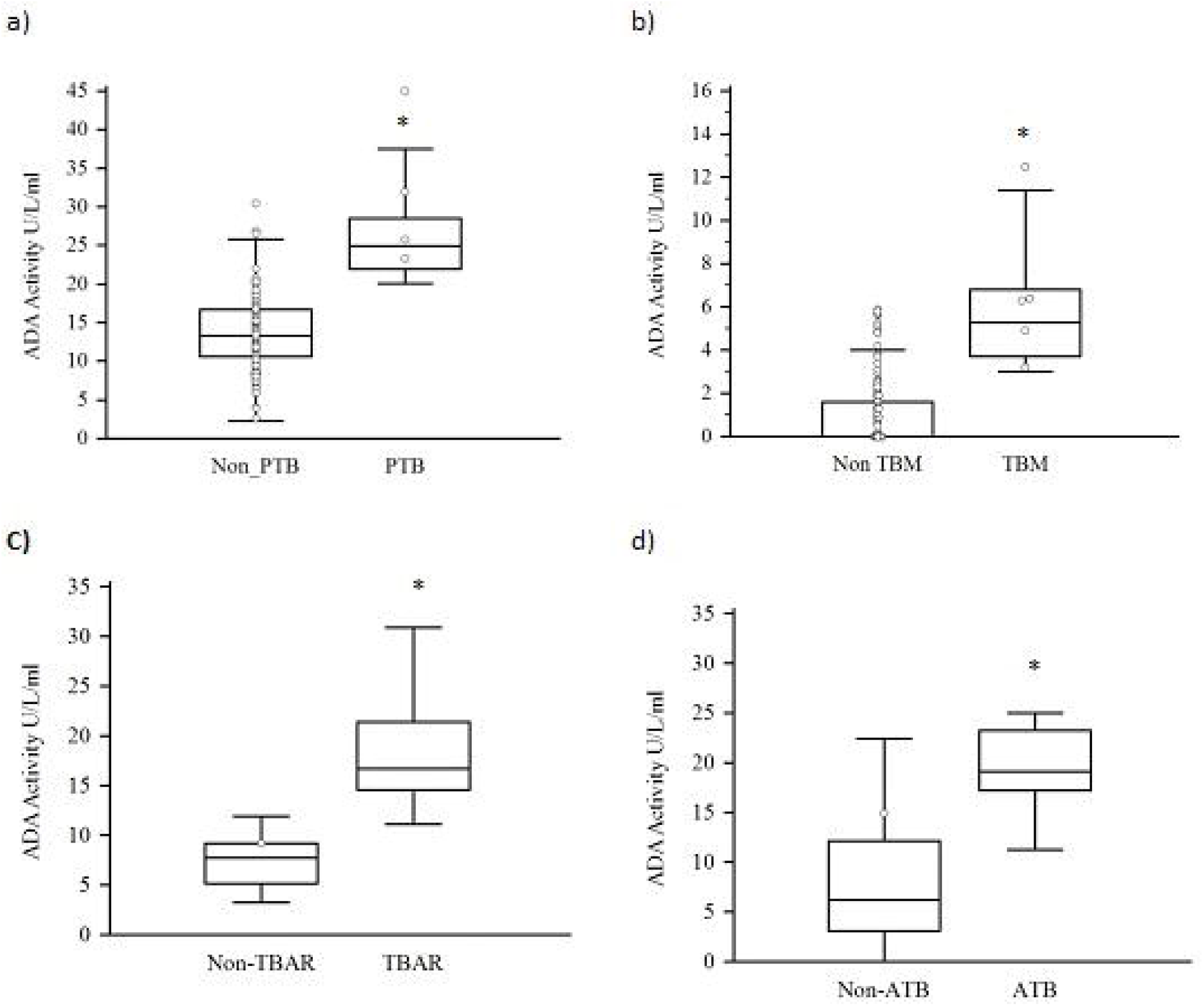
Shows the box plots presentation of ADA activity in PTB, TBM, TBAR, & ATB patients & respective non TB groups. Note: *represent P < 0.05

**Figure 3(a-g):**
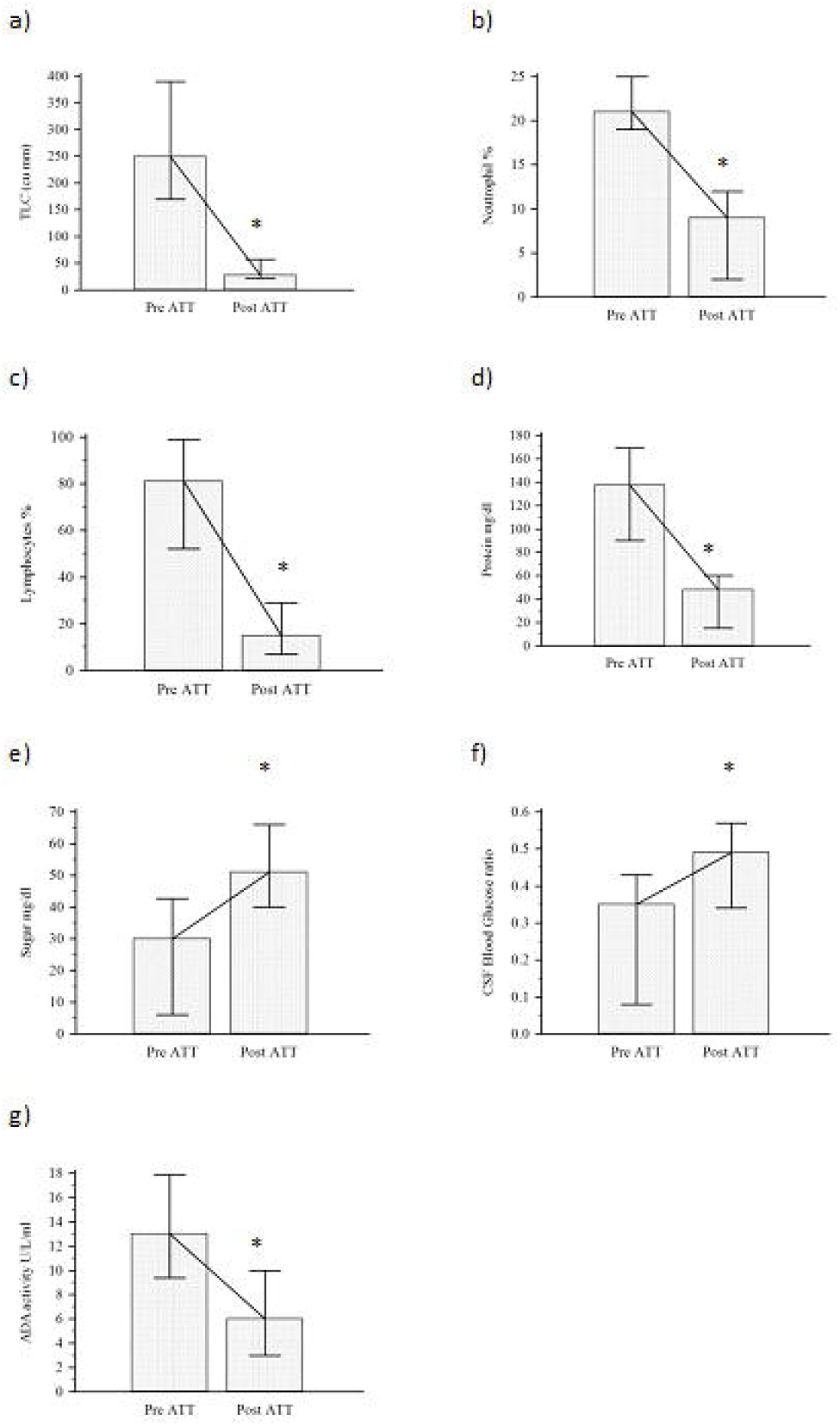
Show the follow-up total leukocyte count (TLC), neutrophil count, Lymphocyte count, Protein, Sugar, CSF:blood glucose ratio and ADA activity (U/dl)] in the CSF samples of TBM patients (n=67) before & after ATT for three months. Note: *represent P < 0.05

**Figure 4(a-c):**
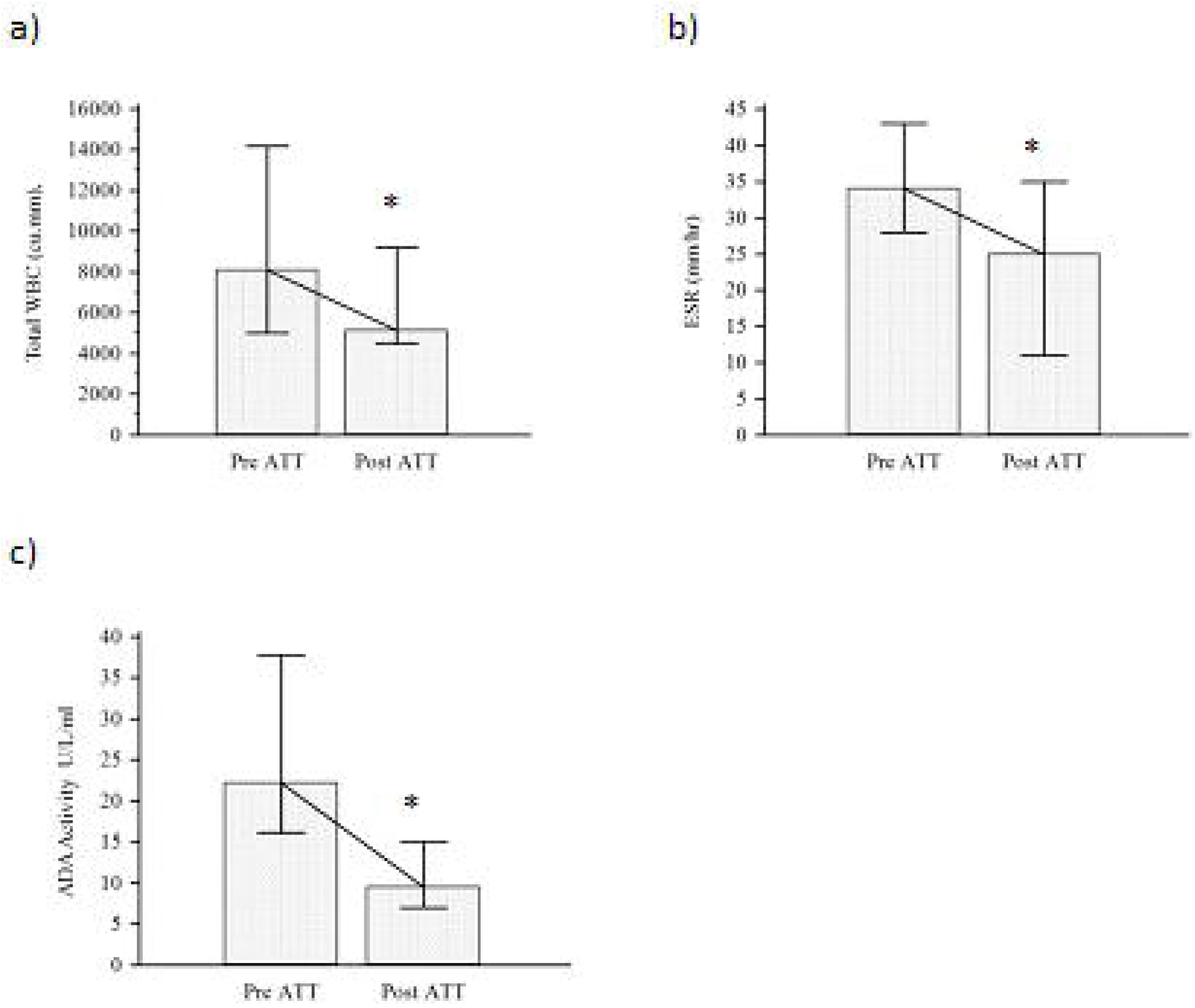
Show the follow-up Total WBC count, Erythrocyte sedimentation rate (ESR) and ADA activity in the serum samples of PTB patients (n=1330) before & after taking ATT for three months. Note: *represent P < 0.05

## 6. Discussion

In the present study, we have assessed the diagnostic value of ADA in serum samples of PTB, and EPTB (TBM [CSF sample], TBAR [Synovial fluid]) & ATB [pleural fluid,ascitic fluid, abdominal fluid] patients. This ADA evaluation study was done in a large number of patients, especially in TBM and PTB cases. We have indeed included a good number of samples of ATB as compared to previous studies to the best of our knowledge. ADA in TBAR is also not much used in earlier studies and due to this, not many cases are reported. We also faced difficulty in getting synovial fluid samples, but as compared to other reports, the number of samples included in the present study is sufficient to conclude the performance of ADA in TBAR cases.

Our data show that both the sensitivity and specificity of the ADA are better in all forms of EPTB cases. In the TBM subjects, we have shown the sensitivity of 85% and specificity of 89% for the ADA test. Several studies, including ours, have suggested the use of ADA levels for the diagnosis of PTB and EPTB. Gupta et al in a study evaluated ADA in 40 CSF samples and reported 94.73% sensitivity and 90.47% specificity in differentiating TBM from non-TBM [14,15] Previously we have also reported a sensitivity of 82% and specificity of 83% in 281 TBM and non-TBM patients [10]. However, in this study, we have taken a high number of samples (n=7498) as compared to previous studies, but the sensitivity and specificity are more or less similar to earlier studies [7]. Agrawal et al studied ADA levels in 56 patients and obtained a sensitivity of 87.5 % and specificity of 88.33 % [16,17]. All these studies concluded that ADA estimation is a fairly specific method for making a diagnosis of tuberculous etiology in TBM, especially when there is a dilemma of differentiating the tuberculous etiology from non-tuberculous ones. Despite of it there is a debate on the use of the ADA test for diagnosis of TBM. In the present study, however, to overcome the above-mentioned problems related to sample numbers, we have included a large number of samples and the results are encouraging which suggests that the ADA is the best option to use for diagnosis of TBM. Similarly, the use of the ADA in ATB is also not much clear as per the previously published work. The data, we have reported in the present study suggests that the diagnosis of PTB using pleural fluid by ADA assay becomes an efficacious and inexpensive method and seems to have good diagnostic accuracy.

TBAR is another form of tuberculosis, which was not investigated in detail. Only a few numbers of have been conducted regarding the diagnostic value of ADA in synovial fluid [18,19]. In the present study, we have reported good sensitivity and specificity of the ADA test for the diagnosis of TBAR. The sample number of this group is very low, as we have received only a few cases during the study period. The results obtained suggest that it can be one of the best cost-effective markers for the rapid diagnosis of TBAR. A similar observation was reported by Chingching Foocharoen et al, as they have shown that Synovial fluid ADA levels in non-TB septic arthritis patients, was higher than in other types of inflammatory arthritis [18]. As mentioned elsewhere not many studies done on the ADA with reference to TBAR, however, in another study done by R Kumar et al showed the elevated value of ADA activity was observed in the synovial fluid of patients with TBAR. Thus, we recommend that the estimation of ADA activity in synovial fluid can be used for the diagnosis of TBAR [19].

Similar to our findings, numerous studies conducted in different fluids of ATB patients have reported the diagnostic utility of ADA assay. Valdes et al demonstrated that the activity of both isoenzymes contributes to the high ADA activity in tuberculous pleural fluids, with ADA-2 playing the predominant role [20]. Khan et al have reported that the sensitivity of an elevated level of ADA varies between 56% and 100%, while the specificity ranges from 55% to 100% [21]. The sensitivity and specificity of ADA in our study were observed to have fallen within the above-mentioned range using a good number of samples. This study therefore clearly suggests that the ADA can be a useful and reliable marker for diagnosing ATB patients.

Despite of getting encouraging results for ADA as a diagnostic marker for EPTB, especially TBM, TBAR, and ATB, unfortunately, we have not got similar results for PTB. In our study concerning this group, we have shown good sensitivity with the ADA test (82%) but very poor specificity (48%). In our study, we have found increased levels of ADA in serum due to several causes, including other bacterial infections, rheumatic disease, and lymphoproliferative disorders [7, 22]. The possible reason for the poor specificity of the ADA in serum samples of PTB as compared to EPTB cases (especially in CSF, synovial fluid) is the load of protein sample. The protein in serum is 6-7gm /dL as compared to 30-34mg/dL in CSF and possibly due to the high load of protein in serum, which contains various host proteins, external antigens which may produce their specific T cell lymphocyte even when TB antigens are absent. As we are aware that ADA is an enzyme that increases during TB infection because of the stimulation of T-cell lymphocytes by mycobacterial antigens, but in serum, it may increase due to the presence of other several antigens and host proteins produced in other infections. This is not the case with CSF and synovial fluid, which are sterile body fluids as compared to serum. Many studies suggest that despite good accuracy, ADA activity has poor specificity and quite variable sensitivity in detecting PTB. The second common problem encountered with the ADA test is that it does not have a defined and universal cut-off value which can be used for calculating sensitivity and specificity [10, 22,23,24]. Lastly, some mutations in the gene of ADA cause it to be unexpressed [25,26]. Deficient levels of ADA have also been associated with pulmonary inflammation, thymic cell death, and defective T-cell receptor signaling [27,28]. These observations indicate that neither a positive value for ADA activity in serum samples confirm the diagnosis of PTB nor negative value of ADA, rules out the presence of PTB infection. Thus, ADA tests have little role in the diagnosis of PTB.

Other than the diagnostic role of the ADA in PTB and EPTB cases, we also evaluated whether ADA can be useful as a prognostic marker for all forms of TB. Our results suggest that ADA might have prognostic potential in TB cases. In the follow-up studies carried on in PTB and TBM patients, we observed a sequential decrease in ADA levels in patients, who took anti-Tb treatment (ATT) for three months. In TBM subjects, the ADA was also compared with routine CSF parameters like TLC, protein, sugar, etc, used routinely for the diagnosis and prognosis. ADA level declined with treatment along with mentioned CSF parameters. Perhaps this decrease might reflect the normalization of the altered lymphocyte turnover induced by TB. The measurement of ADA activity could be of some help to evaluate the response to ATT in PTB and TBM patients. Earlier few studies have suggested ADA as a prognostic indicator and shown a decrease in ADA levels with ATT [29,30].

## 7. Conclusion

In the present study, using a large number of clinical samples of PTB and EPTB cases, we have concluded that the use of ADA test in diagnosing EPTB cases like TBM, TBAR & ATB is useful and can be used for diagnosis in the light of the patient’s clinical condition. On the other hand, our study suggests that ADA has very limited value in the diagnosis of PTB. In addition to that, our follow-up study results, suggests that ADA will be useful as a prognostic indicator for all forms of TB.

## Data Availability

All the data was generated during the course of the study.

## 8. Acknowledgements

All authors would like to acknowledge Dr. G.M Taori Central India Institute of Medical Sciences (CIIMS) for funding this study. We would also like to acknowledge Mr. Ajaz S. Ali, Ms. Sonali Gaikwad, Ms. Ruchika Jain, Ms. Shweta Deote, Mr. Hari Gaherwar and Mrs. Sonali Ramteke for their technical assistance.

## 10. Authors’ contributions

Study design (RSK, HFD, NCC), Data collection (ARM, VRH, SDS), Statistical analysis & data interpretation (ARN, RNB), Literature search (RSK, ANM, ARN, RNB), Manuscript preparation (RSK, ANM, ARN), Assisted in data analysis collection (RSK, ARN, NCC, HFD, VSA), Data interpretation (RSK, ARN, ANM), Participated in the preparation of the manuscript (RSK, ANM), Provided assistance in preparation of the manuscript, data interpretation, study design, and funds collection (RNB, ARN, VSA, LRS), Supervised the study design and statistical analysis (RSK & ARN), Data interpretation, manuscript preparation, and literature search (RNB, ANM).

